# Improved Multiple Sclerosis Lesion Detection using an Intelligent Automation Software

**DOI:** 10.1101/2022.06.22.22276781

**Authors:** Christian Federau, Nicolin Hainc, Myriam Edjlali, Guangming Zhu, Milica Mastilovic, Nathalie Nierobisch, Jan-Philipp Uhlemann, Silvio Paganucci, Cristina Granziera, Olivier Heinzlef, Lucas B. Kipp, Max Wintermark

**Author notes:** Corresponding Author: Christian Federau, AI Medical AG, Goldhaldenstr 22a, 8702 Zollikon, Switzerland.

## Abstract

**Introduction:** The assessment of multiple sclerosis (MS) lesions on follow-up magnetic resonance imaging (MRI) is tedious, time-consuming, and error-prone. Automation of low-level tasks could enhance the radiologist in this work. We evaluate the intelligent automation software Jazz in a blinded three centers study, for the assessment of new, slowly expanding, and contrast-enhancing MS lesions

**Methods:** In three separate centers, 117 MS follow-up MRIs were blindly analyzed on FLuid Attenuated Inversion Recovery (FLAIR), pre- and post-Gadolinium T1-weighted images using Jazz by 2 neuroradiologists in each center. The reading time was recorded. The ground truth was defined in a second reading by side-by-side comparison of both reports from Jazz and the standard clinical report. The number of described new, slowly expanding, and contrast-enhancing lesions described with Jazz was compared to the lesions described in the standard clinical report.

**Results:** A total of 96 new lesions from 41 patients and 162 slowly expanding lesions (SELs) from 61 patients were described in the ground truth reading. A significantly larger number of new lesions were described using Jazz compared to the standard clinical report (63 versus 24). No SELs were reported in the standard clinical report, while 95 SELs were reported on average using Jazz. A total of 4 new contrast-enhancing lesions were found in all reports. The reading with Jazz was very time efficient, taking on average 2min33sec ± 1min0sec per case. Overall inter-reader agreement for new lesions between the readers using Jazz was moderate for new lesions (Cohen kappa=0.5) and slight for SELs (0.08).

**Discussion:** The quality and the productivity of neuroradiological reading of MS follow-up MRI scans can be significantly improved using a dedicated software such as Jazz.

## Introduction

Multiple sclerosis (MS) is an autoimmune, potentially disabling demyelinating disease of the central nervous system (CNS), with clinical onset typically in the young adult. ^1^ Traditionally, MS progression have been categorized in three main type, Primary-Progressive MS (PPMS) affecting between 10 and 15 % of MS patients, Relapsing-Remitting MS (RRMS) and Secondary-Progressive MS (SPMS), ^1^ although it has been recently suggested that the clinical course of MS is better described as a continuum, with contributions from concurrent pathophysiological processes that vary across individuals and over time. ^2^ Prior to the availability of the disease-modifying therapies, roughly 50% of patients diagnosed with RRMS would transition to SPMS within 10 years, and 90% would transition within 25 years, with a median time of about 19 years. ^3^ The diagnosis of SPMS is most often established retrospectively, ^4^ on average 3 years after the actual progression started. ^5^

This is caused by the difficulties by the clinicians and the patients to interpret the initial symptoms indicating early progression, which they can be subtle and fluctuating, ^4^ and by the difficulties to detect disease progression on imaging. The importance of detecting remaining inflammatory disease earlier and more systematically has further increased with the demonstration that early intervention with high-efficacy disease modifying therapies can delay the onset of SPMS, ^6^ and with the recent availability of approved new therapies for progressive MS, such as the anti-CD20 monoclonal antibody Ocrelizumab ^7^ or the selective sphingosine-1-phosphate receptor modulator Siponimod. ^8^

MS disease monitoring with imaging is particularly challenging. It involves at least yearly follow-up magnetic resonance imaging (MRI) scans of their central nervous system, and consists of the tedious, time-consuming, and error-prone manual comparison and counting of the multiple demyelinating lesions. Given time constraints and the fact lesion burden can reach hundreds in severe cases, neuroradiologists can be forced to grossly compare the lesion load, and relevant disease evolution might remain unnoticed for some time. Longitudinal subtraction maps have been used to increase sensitivity for detecting new or enlarged lesions multiple sclerosis lesions^9–12^ and reduce reading time^13^, but small lesions might be difficult to detect using this technique.^14^

Fully automated neuroradiological reporting of MS lesion load evolution would be of great interest, but current methods, including deep learning methods, have currently limited practicality given they are far from error free. In the last decade, deep learning methods, in particular two-dimensional and three-dimensional convolutional neural networks, have been applied to develop automatic lesion detection and segmentation. ^15^ Reported lesion-wise true positive rate ranges between 0.15 and 0.57, and the lesion-wise false positive rate ranges between 0.08 and 0.68. ^16–25^ Those numbers are relatively low, given that every single new or evolving lesion on a follow-up control can be considered a marker of disease progression requiring a re-evaluation of the patient’s current therapy, and therefore, the expert radiologist assessment is still required for every patient. ^26^

In this context, a tool based on intelligent automation, which could simplify the work of the radiologist by automating many repetitive tasks, would be of interest. Intelligent automation aims to streamline processes to improve efficiency and reduce errors. ^27^ It applies the concept of breaking larger complex tasks into several simpler, very-well-understood low-level steps, and automatizes the low-level steps with intelligent software, using artificial intelligence and advanced software engineering. By freeing the radiologist to concentrate on the lesion assessment, it could reduce the number of missed lesions. Intelligent automation promises to improve productivity, reduce costs, as well as more precise processes and a better user experience. Jazz ^28^ is an intelligent automation software dedicated for neuroradiology, that automates multiple low-level tasks such as contrast recognition, previous exams organization, and images coregistration. It includes a semi-automatic lesion annotation tool, permitting the user to easily save all relevant information in a lesion list, and has a graphical user interface intended to maximize productivity and to minimize operator fatigue and discomfort. (**Figure 1)** In this retrospective study, we evaluate Jazz for the assessment of new, expanding, and contrast-enhancing MS lesions, blindly in three centers, and compared the reported lesions with the lesions described in the standard clinical report.

**1.**
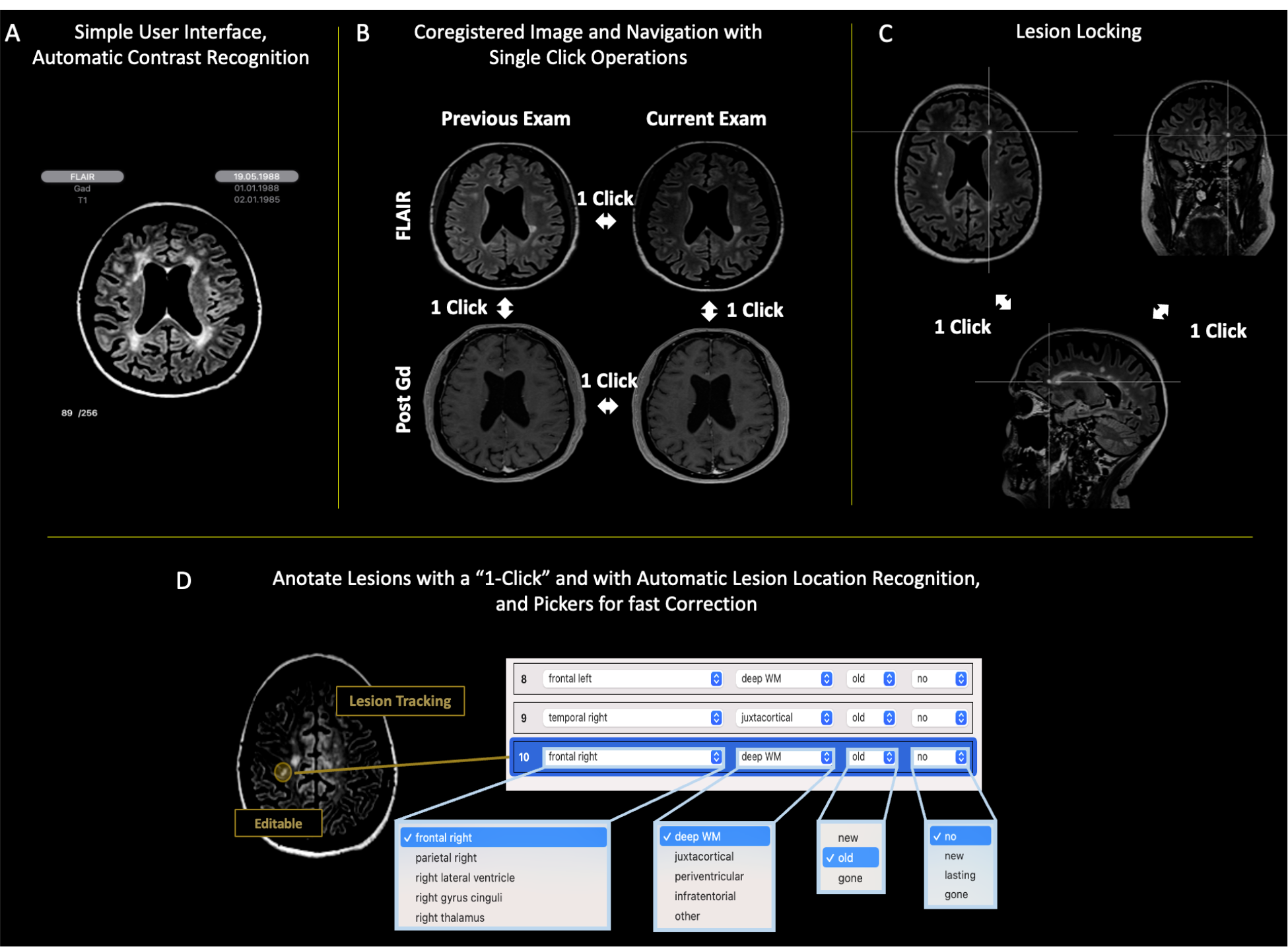
The intelligent automation software automizes low-level tasks for the radiologists, including (**A**) automatic contrast recognition, exams ordering, and (**B**) image coregistration for fast “single-click” lesion comparison and evaluation “without eye movements”, maximizing efficiency and minimizing errors. (**C**) Lesion locking, which permits the evaluation of the lesion in various plane with a “single-click”. (**D**) Automatic anatomic localization and lesion tracking permits ultra-fast lesion navigation. The pre-populated drop-down pickers allow for fast corrections of lesion characteristics. From the lesion list, the software counts the lesions, summarizes the findings, and generates a report automatically, which permits an additional gain in time. Automated lesion counting and generation of a standardized, editable report, including overview figures of the lesions, saves additional time for the radiologist, and eliminates possible human errors in this step.

## Methods

The following analysis was performed in three separate centers in a fully blinded and independent manner.

### Dataset

A set of 120 (40 from each of the 3 centers) current and prior magnetic resonance examinations of patients diagnosed with MS were obtained. 3D FLAIR, pre- and post-Gadolinium T1-weighted 3D sequences, as well as the corresponding clinical report, were analyzed. (**Table 1**) All data were anonymized.

**Table 1.** Number of patients with true positive new lesions.

|  | **Ground**<br>**Truth** | **Jazz**<br>**Reader 1** | **Jazz**<br>**Reader 2** | **Jazz Reader**<br>**Average** | **Standard**<br>**Clinical**<br>**Report** |
| --- | --- | --- | --- | --- | --- |
| Center 1 | 12 | 5 | 12 | 8.5 | 3 |
| Center 2 | 11 | 9 | 10 | 9.5 | 4 |
| Center 3 | 18 | 13 | 12 | 12.5 | 6 |
| **Total** | **41** | **27** | **34** | **30.5** | **13** |

### Case Reading using Jazz

The cases were loaded in Jazz. In a pre-processing step, Jazz automatically coregistered the images to one another using an affine transformation and labelled their contrast. The reader used then Jazz user interface to switch between the coregistered exams and contrasts using single-click operations or using the corresponding keyboard short-cut to evaluate the lesions. If necessary, the reader used the lesion locking tool to evaluate the lesion in different planes. When the user detected a new, SEL, or contrast-enhancing lesion, the reader used Jazz lesion annotation tool, which permits marking of observed lesions with a single click, wherein the software automatically recognized the anatomical lesion location, and used if necessary the interactive lesion list with picker menus to correct the lesion description if necessary. (**Figure 1**)

In each center, two neuroradiologists evaluated independently all 40 cases using the Jazz software (center 1 __Blinded for Review__, 12 years of experience, and __ Blinded for Review__, 8 years of experience, center 2, __ Blinded for Review__, 15 years of experience, and Blinded for Review__, 2 years of experience, center 3 __ Blinded for Review__, 5 years of experience, and __ Blinded for Review__, 3 years of experience). The new lesions and the slowly expanding lesions (SELs) were assessed on the 3D FLAIR. New lesions were defined as non-preexisting white matter lesions on the previous exam, visible on the current exam, while SELs were defined as preexisting white matter lesions that demonstrate lesions expansion between previous and current exam. The contrast-enhancing lesions were assessed by comparing the pre- and post-GAD T1-weighted 3D sequences. Reading time was recorded.

### Ground Truth

A ground truth was defined by reevaluating, using side-by-side comparison, all lesions described in the standard clinical reports and the reports generated by Jazz. The ground truth reading was done by either the most experienced reader of both readers (in two centers), or by a third experienced reader (third center, __ Blinded for Review__, 8 years of experience). Reported new lesions, SELs, and contrast-enhancing lesions in each report were then counted, and classified as true positive, false negative, or false positive.

### Statistical Analysis

Statistical significance was assessed using the Mann-Whitney U test between the average reading time with Jazz and generating the standard report. Significance level was set to α < 0.05. Inter-reader agreement between the readers using Jazz was assessed using Cohen’s Kappa.

## Results

### Patient Population

39 cases were included from center 1 (mean age 55.5 ± 10.6 y, median age 57 y, age range 25-72 y, female / male = 30 / 9; one case was excluded because the lesions were very confluent, rendering the comparison of separate lesions difficult). 40 cases were included from center 2 (mean age 48.3 ± 12.8 y, median age 45 y, age range 26-73 y, female / male = 31 / 9). 38 cases were included from center 3 (mean age 35.4 ± 9.8 y, median age 34 y, age range 20-64 y, female / male = 20 / 18; one case was excluded because the follow-up MRI was done for hydrocephalus evaluation without dedicated description of the MS lesions, and one case was excluded because the 3D FLAIR was missing in one of the exams).

### Reading time

Reading time using Jazz took on average 2 min 33 sec ± 1 min 0 sec per case for all readers (**Figure 2**). In center 1, reading using Jazz took 2 min 29 sec ± 2 min 15 sec per case for reader 1, and 1 min 45 sec ± 51 sec for reader 2. In center 2, reading using Jazz took 4 min 30 sec ± 2 min 42 sec per case for reader 1, and 2 min 33 sec ± 1 min 12 sec for reader 2. In center 3, reading using Jazz took 2 min 12 sec ± 1 min 7 sec per case for reader 1, and 1 min 51 sec ± 1 min 4 sec for reader 2.

**2.**
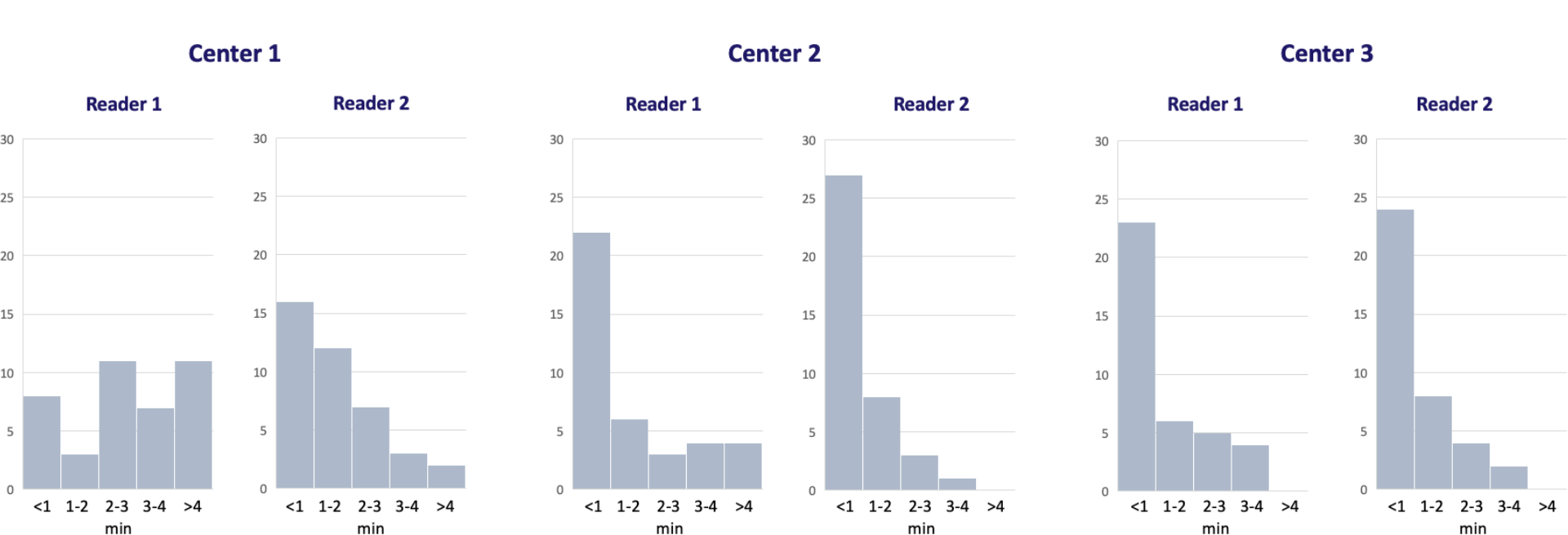
Histogram of the reading time by each reader in the three centers using Jazz.

### New Lesions on 3D FLAIR

In all three centers, the ground truth reading reported an average of 0.83 ± 0.08 new lesions per case (a total of 96 lesions from 41 patients, out of a total of 117 patients). The standard clinical reports reported 24 true positive new lesions from 13 patients, 72 false negative, and 5 false positive new lesions. The Jazz report reported on average (average of 2 readers) 63 true positive lesions from 30.5 patients, 33 false negative, and 18 false positive new lesions (p < 0.05; **Table 1 and Figure 3**).

**3.**
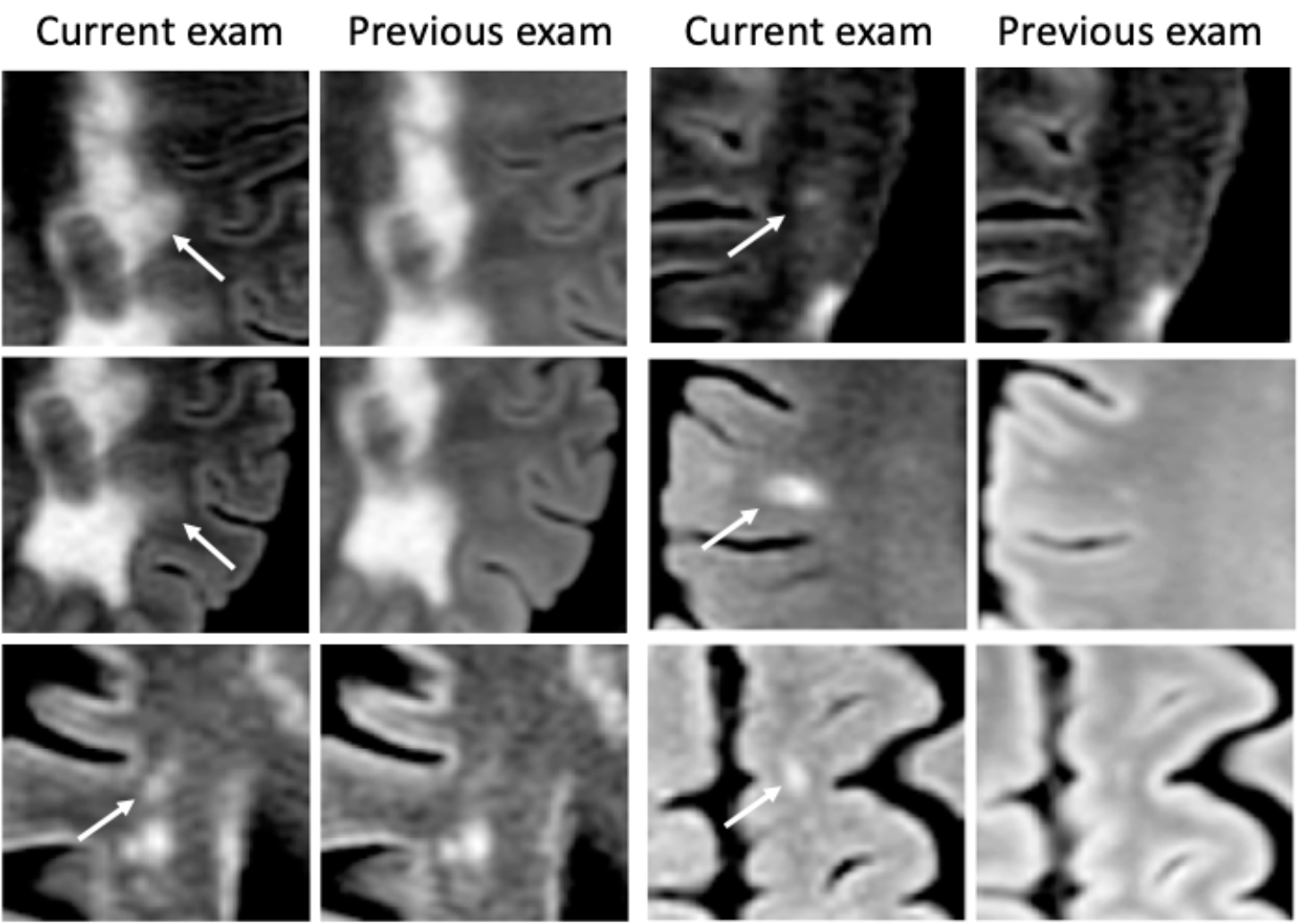
Examples of missed lesions on the standard report, detected by both readers with the Jazz software.

### Slowly Expanding Lesions on 3D FLAIR

In all three centers, the ground truth reading reported an average of 1.39 ± 0.72 SELs per case (a total of 162 lesions from 61 patients, out of a total of 117 patients). The standard clinical reports reported no growing lesions at any center. The Jazz reports reported on average (average of 2 readers) 95 true positive growing lesions from 45 patients, 67 false negative, and 38.5 false positive SELs (p < 0.05; **Table 2 and Figure 4**).

**Table 2.**
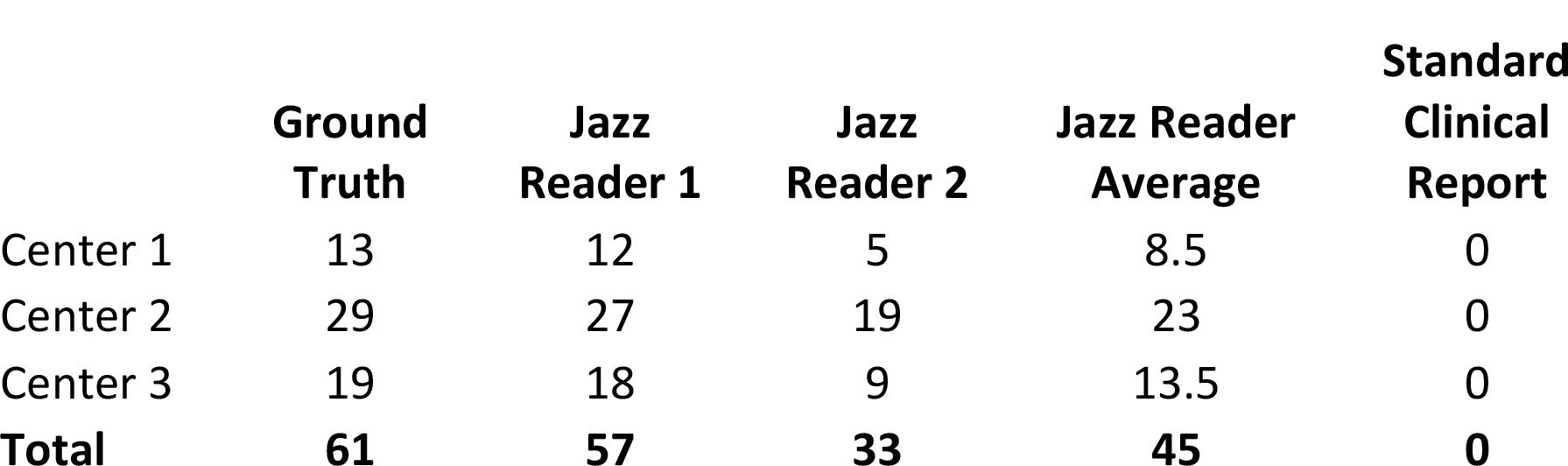
Number of patients with true positive SELs.

**4.**
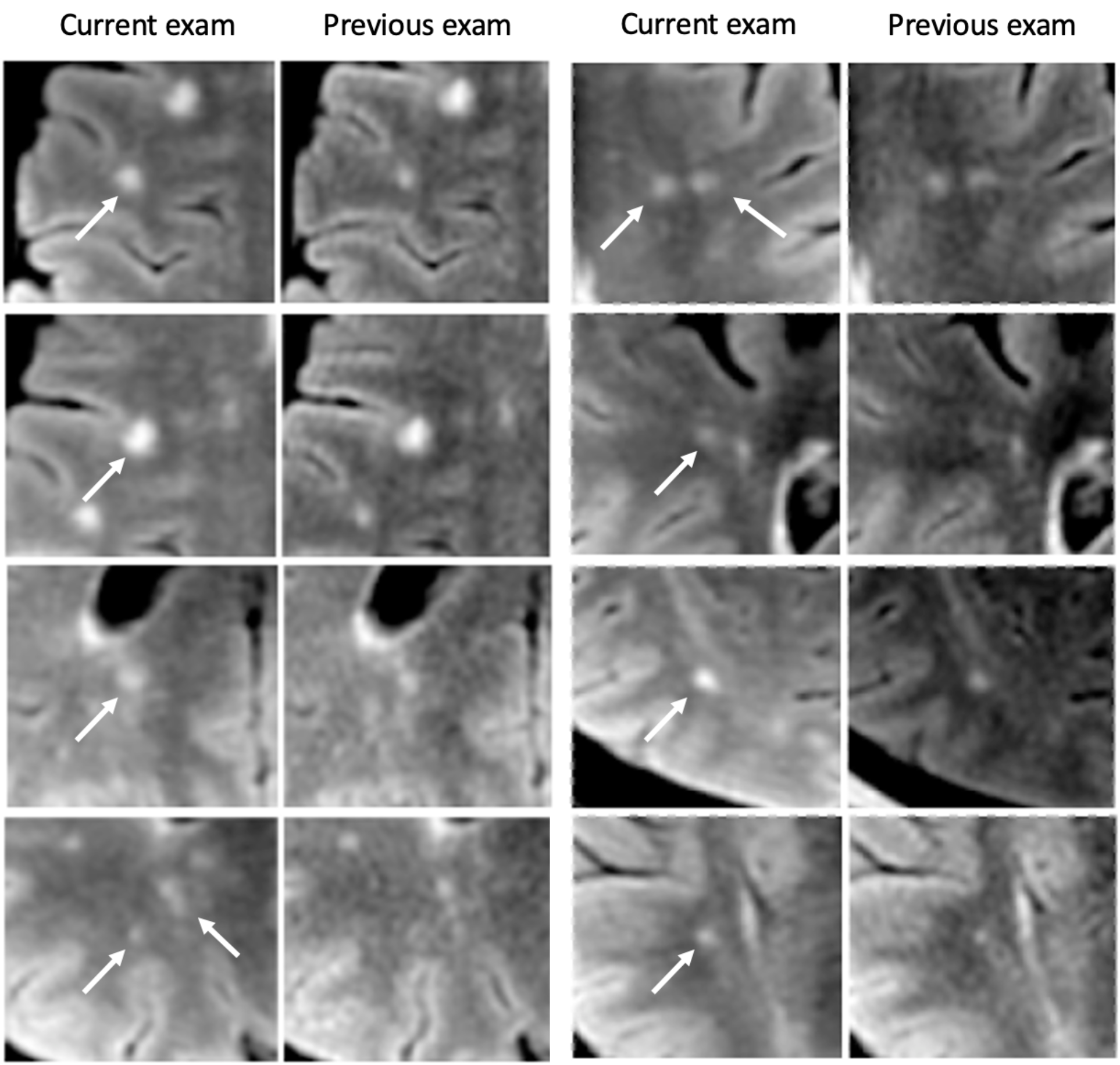
Examples of SELs not described on the standard clinical report, but described by both readers with the Jazz software. Note that all those images were acquired from the same patient. While the presence of a single SEL might be often difficult to interpret, as it might occur from partial volume effects or from MRI artifacts, the presence of multiples SELs is highly suggestive of disease progression.

### Contrast-enhancing Lesions

In center 1, all patients were administered intravenous contrast, and no contrast-enhancing lesions were described in any report. In center 2, 11 patients were not administered intravenous contrast; in the remaining 29 patients, 4 new contrast-enhancing lesions were described, consistently in all reports, and 1 false positive contrast-enhancing lesion was described on the standard clinical report only. In center 3, all patients except one were administered intravenous contrast; one contrast-enhancing lesion was described, consistently in all reports.

### Inter-reader Agreement

Overall inter-reader agreement for new lesions between the readers using Jazz was moderate, with kappa = 0.5 (center 1: 0.45, center 2: 0.51, center 3: 0.54) and for SELs it was slight, with kappa = 0.08 (center 1: -0.3, center 2: 0.3, center 3: 0.22).

### Statement

This study was approved by the Ethic committee of the Université Paris, of the Canton of Zürich (2022-00041) and of Stanford University (IRB-26147). All methods were carried out in accordance with relevant guidelines and regulations. Informed consent from the subjects was waived by the above-mentioned ethic committee. The datasets used and analyzed during the current study available from the corresponding author on reasonable request.

## Discussion

This study shows that a significantly larger number of new and SELs are reported when using the intelligent automation software Jazz compared to the standard clinical reporting method in a time efficient manner, by allowing the neuroradiologist to concentrate on the detection and evaluation of lesions. By permitting to toggle between coregistered current and previous exams, it uses a particularly potent neurophysiological mechanism to attract the attention of the reader to modifications in the images, in other words in the case of MS, to new and evolving lesions. Indeed, attention is a selective process and is physiologically necessary, because there are severe limits on our capacity to process visual information. ^29^ A theory suggests that attention is imposed by the fixed amount of overall energy available to the brain, and that because of the high-energy cost of neuronal activity, only a small fraction of the machinery can be engaged concurrently. ^29^

As a consequence, stimuli have to compete for limited resources, ^30^ a notion that is supported by electrophysiological, neuroimaging and behavioral studies. ^31,32^ Of the several visual stimuli that are known to capture attention, the sudden appearance of a new object and the sudden changes of an object are known to be particularly potent and to influence priority in visual search. ^33–35^ This mechanism is exploited in the dynamic switching tool of Jazz, compared to a standard side-by-side comparison method.

The identification of progression in MS is typically retrospective and based on clinical symptoms. ^36^ Most clinical trials rely on an increase in Expanded Disability Status Scale score confirmed over 3 or 6 months to define disease progression, and might overestimate the accumulation of permanent disability by up to 30%. ^37^ Given the profound burden of progressive MS and the recent development of effective treatments for these patients, there is a need to establish reliable and reproducible methods for identifying progression early in the disease course. Neuroradiological MRI reports of MS are often sparse and not standardized, despite the advantages of standardization of clinical data for MS monitoring being well recognized. ^38^ Structured reports of MRI in patients with MS have been shown to provide more adequate information for clinical decision making than nonstructured reports. ^39^ Improvement in the digitization of real-world patient data could foster new insights into the epidemiology and pathophysiology of the disease, and are ultimately necessary to achieve truthful personalized patient care. ^40^

This study also shows that at least one SEL was described in more than half of all patients, while none were reported in the standard clinical reports. The inter-reader agreement was low though, showing that this assessment remains particularly difficult and prone to subjectivity. The use of objective criteria defining SELs might help to improve this aspect. SELs can be easily overlooked, and are particularly difficult to detect in a standard DICOM viewer. There is substantial interest in SELs as a potential marker of chronic but active MS lesions ^41–43^ which may have diagnostic, prognostic and treatment implications. SELs were found to be more prevalent in patients with primary progressive MS (PPMS) compared with patients with relapsing-remitting MS (RRMS). ^42^

The proportion of SELs and their microstructural tissue abnormalities were associated with a higher risk of MS progression and secondary progressive MS (SPMS) conversion. ^44^ In SPMS, SELs were found to represent almost one-third of T2 lesions, associated with neurodegenerative MRI markers and related to clinical worsening. ^45^ Therefore the number and volume of SELs are a promising biomarker to predict a more active, progressive disease course and could become a new target for therapeutic intervention. This is particularly interesting, because recent advances in our understanding of the mechanisms that drive SELs have fueled optimism for improved treatment of this condition. ^46^ A robust tool to gather SELs on magnetic resonance images, such as the one described in this work, might help to detect patients with progressive disease earlier and more systematically.

This study suffers from several limitations. Only three centers were included, and our results may not generalize to other centers. The reading using the Jazz software was not under identical conditions to the reading employed to generate the standard report, which was done during the standard clinical routine and might have included more distracting events. The reading of the standard clinical reports included at least a first reading and a second reading through a supervisor, and discussion with, and comments from the referring clinicians might have occurred. The reading with the Jazz software only included a single reading from a single reader. The ground truth was not based on a consensus reading performed by the most experienced reader in each center, which might have biased the agreement. Current and previous exams could have been acquired on different scanners, which might be a cofounder in the SEL evaluation. Further, a previous definition of SELs was based on the inclusion of a least three time points over more than two years, ^42^ with local expansion assessed by the Jacobian determinant of the deformation between reference and follow-up scans, and a heuristic scored was used to favor individual SEL candidates undergoing concentric and constant change, ^42^ while here, a more pragmatic approach was considered, using the subjective expansion assessment based on only two MR time points. Finally, no standardized method was used to define the SELs, as the evaluation was left to the subjective appraisal of each reader.

In conclusion, this study shows that significantly more new lesions and SELs can be described in a time-efficient manner using the intelligent automation software Jazz compared to the standard method.

## Data Availability

All data produced in the present study are available upon reasonable request to the authors

## Data Statement

The data that support the findings of this study are available from the corresponding author, C.F., on reasonable request.

